# Impacts on Surgery Resident Education at a first wave COVID-19 epicenter

**DOI:** 10.1101/2020.08.16.20176073

**Authors:** Alexander Ostapenko, Samantha McPeck, Shawn Liechty, Daniel Kleiner

**Affiliations:** Department of General Surgery, Danbury Hospital, Danbury CT; Department of Internal Medicine, University of Connecticut, Storrs CT

**Keywords:** General Surgery, Resident education, COVID-19, training

## Abstract

**Background:** This study aims to identify the effects of the COVID-19 pandemic on surgical resident training and education at Danbury Hospital.

**Methods:** We conducted an observational study at a Western Connecticut hospital heavily affected by the first wave of the COVID-19 pandemic to assess its effects on surgical residents, focusing on surgical education, clinical experience, and operative skills development. Objective data was available through recorded work hours, case logs, and formal didactics. In addition, we created an anonymous survey to assess resident perception of their residency experience during the pandemic.

**Results:** There are 22 surgical residents at our institution; all were included in the study. Resident weekly duty hours decreased by 23.9 hours with the majority of clinical time redirected to caring for COVID-19 patients. Independent studying increased by 1.6 hours (26.2%) while weekly didactics decreased by 2.1 hours (35.6%). The operative volume per resident decreased by 65.7% from 35.0 to 12.0 cases for the period of interest, with a disproportionately high effect on junior residents, who experienced a 76.2% decrease. Unsurprisingly, 70% of residents reported a negative effect of the pandemic on their surgical skills.

**Conclusions:** During the first wave of the COVID-19 pandemic, surgical residents’ usual workflows changed dramatically, as much of their time was dedicated to the critical care of patients with COVID-19. However, the consequent opportunity cost was to surgery-specific training; there was a significant decrease in operative cases and time spent in surgical didactics, along with elevated concern about overall preparedness for their intended career.

## Introduction

The COVID-19 pandemic has had a multitude of unprecedented effects on healthcare systems across the United States. Danbury Hospital is located in Western Connecticut, an area considered to be one of the epicenters of the first wave of the pandemic from March-May of 2020. Within the healthcare system, alterations to usual operations and changes in the allocation of human and material resources dramatically changed everyday workflow in hospitals.^1,2,3^ Examples of this include the cancellation of non-emergent operations, the transition of various sectors to Telehealth medicine, and the redistribution of surgical residents to non-surgical services. Other effects on surgical residents have involved modification of duty hour limitations and adjustments to resident education. Around the world, institutions have transitioned to virtual platforms for academic sessions. Many local and national research conferences have been postponed or cancelled, and requirements for standardized exams have changed.^4^ The effects of these revisions in the long term are challenging to predict, particularly because the response to these singular events has varied significantly across states, healthcare systems, and hospitals.^5^ We aim to utilize both quantitative and qualitative data to analyze these effects in the short term, and postulate how this may evolve over time. Specifically, we hypothesize that surgical residents are working fewer hours and logging fewer operative cases, and that along with changes in education and academic opportunities, this has led to rising concern regarding preparedness for future surgical careers.

## Methods

Within the Nuvance Health Network, major changes to usual operations due to the COVID-19 pandemic were instituted between March 3^rd^ and May 25^th^, 2020. We therefore chose this as the period of interest. We focused on three components of surgical education: clinical experience, didactic conferences, and operative volume.

For clinical experience, we compared the number of duty hours logged by residents during the period of interest with the same time in 2019, and calculated a gross difference and percent change. To evaluate didactic conferences, we calculated the average weekly hours of scheduled didactic lectures during the same period in 2019 and 2020. Additionally, we developed a survey to assess resident perception of changes in didactic structure during this time. We examined total major surgeries logged by residents through the ACGME portal during the period of interest, compared to 2019, and calculated a gross difference as well as percent change in order to assess operative opportunities.

The survey was administered anonymously through SurveyMonkey**®** to all surgical residents at Danbury Hospital between June 9^th^ and June 26^th^, 2020. For questions regarding hours, we calculated the perceived difference in hours for each individual response. Mean and standard deviation for the change were calculated for each question. The survey also contained five-point Likert questions, for which the percentage of residents who responded positively with “agree” or “strongly agree” was calculated.

Our Institutional Review Board deemed this study exempt (IRB# 2012208337) and waived the need for participant consent.

## Results

There were 18 surgical residents in 2019 and 22 in 2020, respectively.

### Clinical experience

Clinical experience was assessed in two ways: first, we examined the duty hours logged for clinical work and number of outpatient clinics attended, and second, through several anonymous survey questions, completed by 91% of surgical residents. Surgical residents worked an average of 64.7 hours/week before the COVID-19 pandemic, and 40.8 hours/week during the pandemic, a 37.0% decrease (Table 1). A significant portion of the clinical experience for surgical residents was redirected to caring for COVID-19 patients during the pandemic. In the survey, residents reported working between 28–38 (SD = 26.7) hours/week caring for COVID-19 patients (Table 1). In regards to outpatient clinics, in 2019 for the period of interest residents averaged 4.7 clinics, while in 2020 this decreased by 70.2% to 1.4 clinics per resident.

**Table 1:** Clinical Experience.

| **<u>Part A: Quantitative Data</u>** |  |  |  |  |
| --- | --- | --- | --- | --- |
|  | Before pandemic | During pandemic | Difference | Percent change |
| **Weekly hours of logged <u>clinical</u> work per resident** | 64.7 | 40.8 | 23.9 | - 37.0% |
| **Weekly hours carrying for COVID patients\*** | 0(0) | 28.6-38.6 (26.7) | + 28.6-38.6 (26.7) | NA |
| **Number of outpatient clinics attended per resident** | 4.7 | 1.4 | -3.3 | - 70.2% |
| **<u>Part B: Qualitative Data</u>** |  |  |  |  |
|  | **Agree (%)** |  | **Disagree (%)** |  |
| **Concerned about preparedness to become an attending** | 7 (35%) |  | 7 (35%) |  |
| **Concerned about completing graduation requirements** | 3 (15%) |  | 15 (75%) |  |
| **Concerned about requiring an extra year of training** | 0 (0%) |  | 19 (95%) |  |
| **My experience and education has been positively impacted by the pandemic** | 35% |  | 20% |  |
| **I am attending the same amount of outpatient clinic** | 5% |  | 95% |  |

\*The variable was acquired from the survey.

### Educational experience

An average of 5.9 hours/week were spent on organized didactics before COVID-19 (Table 2). This included 3 hours of protected time for resident education based on the SCORE curriculum, morbidity and mortality conference, grand rounds, specialty attending conferences, and trauma review conference. During the pandemic, all conferences were done remotely through teleconferencing, and the average time spent on organized didactics was 3.8 hours/week – a reduction of 35.6% (Table 2). Residents were split on whether the quality of didactics improved, with 35% reporting an improvement, and 35% perceiving a decrease in quality. Despite a significant drop in clinical work of 22.1 hours per week, residents reported an increase in independent studying of only 1.6 hours per week: from 6.1 to 7.7 hours per week (Table 2).

**Table 2:** Educational Experience.

| **<u>Part A: Quantitative Data</u>** |  |  |  |  |
| --- | --- | --- | --- | --- |
|  | Before pandemic | During pandemic | Difference | Percent |
| **Weekly hours of didactics** | 5.9 | 3.8 | 2.1 | - 35.6 % |
| **Weekly hours studying individually\*** | 6.1(3.1) | 7.7(4.7) | + 1.6(4.7) | + 26.2% |
| **<u>Part B: Qualitative Data</u>** |  |  |  |  |
|  | **Agree (%)** |  | **Disagree (%)** |  |
| **The quality of didactics increased during the pandemic** | 7 (35%) |  | 7 (35%) |  |
| **Concerned about preparedness to become an attending** | 7 (35%) |  | 7 (35%) |  |
| **Concerned about completing graduation requirements** | 3 (15%) |  | 15 (75%) |  |
| **Concerned about requiring an extra year of training** | 0 (0%) |  | 19 (95%) |  |
| **My surgical knowledge has been negatively affected by the pandemic** | 7 (35%) |  | 8 (40%) |  |
| **My experience and education has been positively impacted by the pandemic** | 35% |  | 20% |  |

\*The variable was acquired from the survey.

### Operative Experience

From March 3^rd^ to May 25^th^, 2019, residents at Danbury Hospital logged 600 operative cases. For the same period during the COVID-19 pandemic, residents logged 240 cases, a 60% decrease (Table 3). Since the number of residents in each post-graduate year (PGY) position varied from 2019 to 2020 we calculated number of cases per resident. On average there were 35 cases per resident before the pandemic, which decreased by 65.7% to 12.0 cases per resident during the pandemic (Table 3). Junior residents in PGY1, 2, and 3 positions were disproportionally affected during the pandemic, with a 76.2% decrease from 25.2 to 6.0 cases per resident for the study period. Senior residents in PGY4 and PGY5 positions saw a 49.7% decreased in operative cases, from 59.6 to 30.0 cases per resident. Overall, 70% of residents felt their surgical skills have been negatively affected by the pandemic.

**Table 3:** Operative Experience.

| **<u>Part A: Quantitative Data</u>** |  |  |  |  |
| --- | --- | --- | --- | --- |
|  | Before pandemic | During pandemic | Difference | Percent change |
| **Total major cases** | 600 | 240 | 360 | - 60% |
| **Average major cases per resident** | 35.0 | 12.0 | 23.0 | - 65.7% |
| Senior residents (PGY 4/5) | 59.6 | 30.0 | 29.6 | - 49.7% |
| Junior residents (PGY 1,2,3) | 25.2 | 6.0 | 19.2 | - 76.2% |
| **<u>Part B: Qualitative Data</u>** |  |  |  |  |
|  | **Agree (%)** |  | **Disagree (%)** |  |
| **I feel my surgical skills have been negatively affected by the pandemic** | 14 (70%) |  | 3 (15%) |  |

## Discussion

During the first wave of the COVID-19 pandemic, Danbury Hospital was among the first institutions on the East Coast outside of New York City to be significantly impacted. Its surgical training program trains 22 residents. From March 3^rd^ through May 25^th^, the hospital was put on emergency status and restructured to maximize the number of COVID-19 patients that could be hospitalized. During this time, ICU capacity to manage ventilated patients was increased from 20 beds to 90, with subsequent redistribution of space within the hospital, education and deployment of nursing staff to critical care units, and changes to all residency programs within the hospital. One of the new 20-bed ICU pods was placed under the care of surgical residents under the supervision of critical care surgeons and anesthesiologists. Additionally, all elective operations were canceled with the exception of urgent cancer surgeries.^6^ This restructuring affected surgical clinical experience, weekly protected time for didactics, and operative opportunities for surgical residents.

### Clinical experience

This restructuring of the program is similar to that reported by institutions in other highly affected areas.^7^ Unlike this study, prior literature did not address the effect of this restructuring on resident training, but discussed their experience and offered recommendations on how to safely and effectively prepare hospitals and programs.^8,9^ In this study, we demonstrate that surgical residents had a significant contribution to the overall COVID-19 response at Danbury Hospital, with an average resident spending 28–38 hours per week caring for critical COVID-19 patients (Table 1). This contribution resulted in a decrease in clinical duty hours by 35.1%, from 64.7 to 40.8 hours per week, in addition to fewer outpatient clinics attended, and less operative experience. However, surgical residents spent more time in a critical care setting, which the American Board of Surgery (ABS) identifies as a primary component of general surgery training.^10^ Similarly, Meyer et al. argued that surgical critical care is crucial for practicing surgeons to be able to holistically manage ill patients with life-threatening conditions. ABS has no specific requirement for duration of ICU rotations; instead, it requires a log of 25 critically ill patients. Prior studies describe a wide variability in surgical critical care training and fund of knowledge of graduating residents.^11,12^ Therefore, this sudden increase in critical care training is one of beneficial effects on surgical training.^10^

One of the ways to enhance resident clinical experience to supplement the drop in clinical duties and outpatient clinic attendance through resident involvement in telehealth clinics.^13^ At our institution there were several barriers to this proposal, including a lack of infrastructure to transition to telehealth and the uncertainty of the timing of elective surgeries resumption. Both of these factors resulted in low volume of appointments initially; however, as telehealth became more common incorporation of residents became more feasible. More widespread incorporation of resident involvement in telehealth clinics can potentially be an invaluable supplementation to clinical experience.

### Educational Experience

The COVID-19 pandemic changed surgical resident didactics, resulting in a decrease in protected time for academics. This was a surprising finding, given the implementation of video conferencing and noted reduced clinical responsibilities of residents. All conferences at our institution transitioned to video platforms, allowing presenters to share screens from remote locations and facilitating assembly of large groups of peers in a safe manner. Other advantages of video platforms include the ability to record lectures for viewing outside of scheduled time, and increased ease in inviting leading experts and educators from prestigious academic institutions to present and discuss topics within their specialty.

Another surprising finding in our study was that despite a decrease in clinical duties by 17 hours per week, independent studying only increased by only 1.6 hours/ week. Given this significant decrease in clinical hours, we expected a larger increase in time spent studying independently. Several factors may play a role here. Given the timing of the national surgical in-training exam (ABSITE), residents may have felt less pressure to increase their time in independent study. Residents who formerly utilized independent study time to prepare for elective cases naturally would decrease time spent on this endeavor.

### Operative Experience

Perhaps most obviously, the COVID-19 pandemic significantly impacted the operative component of surgical training (Table 3). The cancelation of all elective cases resulted in a 60% reduction in total major cases logged by residents. This disproportionally affected junior residents, who went from 25.2 to 6.0 cases per resident, a 76.2% decrease during the periods of interest. Meanwhile, senior resident cases decreased by 49.7%, from 59.6 to 30.0 cases per resident. The American Board of Surgery (ABS) decreased the number of required operative cases for graduating seniors as a direct consequence of the pandemic.^14^ However, such a dramatic decrease in surgical volume will likely affect residents at all levels moving forward. In the survey, 70% of residents reported that the pandemic has negatively affected their surgical skills and 35% reported concern about preparedness to become an attending.

The long-term impacts of the pandemic remain to be seen, yet surgery residents still have a limited five years to acquire the clinical knowledge and operative experience to become surgeons. ABS deems the requirement to qualify for board certification is 54 weeks of surgical clinical experience and 750 logged procedures in defined categories.^11^ Therefore, the significant drop in operative volume is concerning, as physical skill is fundamental to surgical education. One solution to supplement the growing deficit of operative experience is simulation-based training (SBT). Prior studies demonstrated that surgical residents value the ability of SBT to expose them to new procedures, but conclusions were divided on the best ways to implement SBT within curricula.^15^ Through SBT, residents can improve dexterity and speed in operative maneuvers and enhance their technical skill.^16^ Resident performance can even be assessed by attendings or colleagues observing remotely through videoconferencing. This provides a unique opportunity to progress physical skills while maintaining social distancing, and additionally provides another outlet for independent study while clinical hours are reduced.

High quality surgical videos can also help compensate for diminished operative volume. Although not a tactile exercise, when viewed in a group setting with discussion driven by faculty, these sessions can supplement resident operative education.^13^ Videos can also play a role in flipped classroom models in which pre-recorded lectures are watched prior to conferences, which enhances knowledge acquisition and enriches discussion.

The main limitation of this study was that it was limited to a single surgical residency program. Therefore, the results may not be generalizable to residents in other programs in the United States. However, as one of the earliest areas affected by COVID-19, we are able to analyze its effects in a timely fashion that may benefit other geographic areas affected similarly in the future. While residents in states with lower incidence of COVID-19 may not be as significantly impacted as residents at our institution, continued evolution of the COVID pandemic and the rise of new epicenters of disease may make these results more generalizable over time. Despite the limitations, these results are integral in critically thinking about the future of surgical education. The COVID-19 pandemic will continue to affect residency programs across the country with changes to clinical work, didactics, and operative experience of surgical residents.

As physicians, our highest mandate is patient care. We have an ethical and moral responsibility to take care of COVID-19 patients, and there is a great deal to be learned from such experiences. Nonetheless, the cultivation of surgical knowledge and physical skills is integral to the development of future surgeons, and the short duration of residency education is an incomparably formative time. It is important to keep in mind that the role COVID-19 as a disease will have in the future of medical care is impossible to divine, and that regardless of the role it plays, medicine will still need the specific capabilities for which surgeons are trained.

## Conclusion

During the first wave of the pandemic, surgical residents had a significant contribution to care of patients with COVID-19. The impacts of the pandemic on surgeon training continue to evolve, and undoubtedly will have complex long-term effects, both positive and negative. It is important to continually assess how resident training is affected, and to consider innovative approaches to maintain clinical, operative, and educational experiences.

## Data Availability

All pertinent data is available in the manuscript tables.

